# DNA methylation changes in infants of mothers with SARS-CoV-2 infection during pregnancy

**DOI:** 10.1101/2025.05.10.25327356

**Authors:** Eirini Apostolou, Shumaila Sayyab, Marie Blomberg, Camilla Janefjord, Naomi Yamada-Fower, Hanna Östling, Anna Sand, Helena Backman, Malin Veje, Mehreen Zaigham, Sophia Brismar Wendel, Ylva Carlsson, Magnus Domellöf, Fredrik Ahlsson, Sofie Arnkil, Ola Andersson, Lina Bergman, Anders Rosén, Verena Sengpiel, Maria Lerm, Thomas Abrahamsson

**Author notes:** Corresponding author: Maria Lermm Email address of the corresponding author. Shared first authorship (Eirini Apostolou, Shumaila Sayyab). Shared last authorship (Maria Lerm, Thomas Abrahamsson). Lead contact: Maria Lerm.

## Abstract

Maternal environmental exposures, including viral infections, can exert intergenerational effects on the offspring’s epigenome. Herein, based on the Swedish COVID-19 during Pregnancy and Early childhood study (COPE), we explored how mild SARS-CoV-2 infection during pregnancy affected the DNA methylome of peripheral blood mononuclear cells in infants, using Illumina EPIC array. We found significant alterations in DNA methylation patterns in genes related to neurodevelopmental pathways that overlap with cellular processes hijacked by viruses for their own replication. The alterations were further confirmed in cord blood mononuclear cells. Subsequently, we determined the functional consequences of epigenetic rewiring due to maternal SARS-CoV-2 infection without fetal infection and found that fetal exposure to mediators driven by maternal infection, results in an altered responsiveness of the infant’s immune cells upon subsequent cell activation *in vitro*. Finally, we demonstrate that these functional alterations were linked to the identified DNA methylation changes.

## Introduction

Severe Acute Respiratory Syndrome Coronavirus 2 (SARS-CoV-2) infection induces long-term epigenetic reprogramming in adult immune cells, with implications for subsequent immune responses. For example, monocytes from convalescent individuals exhibit altered chromatin accessibility profiles, consistent with trained immunity.^1^ Furthermore, persistent epigenetic and transcriptional changes in hematopoietic stem and progenitor cells (HSPCs) have been associated with enhanced myelopoiesis and altered monocyte phenotypes, pointing to persistent innate immune alterations.^2^ Epigenetic modifications, particularly DNA methylation (DNAm) changes, are prominent in monocytes and T cells of Coronavirus Disease 2019 (COVID-19) convalescents, irrespective of disease severity.^3^ Even asymptomatic or mild SARS-CoV-2 infections have been shown to induce enduring epigenetic effects, including modulation of pathways affecting odor perception^4^ and persistent alterations in interferon-stimulated genes.^5^

Furthermore, significant progress has been made in understanding epigenetic changes in post-COVID patients ^6–9^ and the observed alterations could be mapped to pathways of relevance for post-COVID-associated symptoms such as blood pressure regulation, odor perception, and mitochondrial function.^6^ Epigenetic alterations are also observed in other post-infectious fatigue syndromes.^10^

While accumulated evidence supports persisting epigenetic changes in response to SARS-CoV-2 infection, little is known about the intergenerational epigenetic effects of gestational SARS-CoV-2 infection on progeny epigenetics. Maternal environmental exposures, including stress, dietary shifts, and infections, are well-documented to exert profound effects on the offspring.^11,12^ While the long-term impact of maternal SARS-CoV-2 infection during pregnancy remains unclear, preliminary studies suggest heightened risks for neurodevelopmental and immune dysregulation in neonates.^13^ Maternal SARS-CoV-2 infection has been shown to transiently elevate inflammatory cytokine levels in cord blood and alter neonatal immune cell cytokine responsiveness. These immune perturbations appear independent of the trimester of maternal infection and involve increased percentage/proportion of innate immune cells in neonates, suggesting potential vertical immune priming.^14^

In this exploratory, nested study, within the COVID-19 in Pregnancy and Early childhood study (COPE), we identified alterations in DNAm patterns in immune cells of infants born to mothers with asymptomatic or mild gestational SARS-CoV-2 infection. By analyzing both cord blood and samples collected from infants during the first months of life, we demonstrated that epigenetic changes correspond to functional alterations in immune cells, as validated by in vitro responses to subsequent triggers. Our findings reveal that maternal SARS-CoV-2 infection induces epigenetic rewiring in offspring, mapping to pathways exploited by the virus. The consequences of this remain unknown, but several ongoing studies, including the COPE study, are elucidating the potential long-term effects of the COVID-19 pandemic on children’s development and disease risks.

## Results

### Study design and characteristics

The present study was a sub-study of the COPE study, which was a nationwide multi-center study enrolling pregnant women in Sweden during the COVID-19 pandemic between June 2020 and December 2022. The COPE study is registered at ClinicalTrials.gov (NCT 04433364) and the study protocol has been published elsewhere.^15^ Mothers with and without confirmed SARS-CoV-2 infection during pregnancy were recruited before birth during antenatal care visits and at the delivery departments. Peripheral blood mononuclear cells (PBMCs) were collected from 38 infants (SARS-CoV2, n=23; Controls, n=15) at 1-2 months of age, while cord blood mononuclear cells (CBMCs) were obtained at birth from a subset of dyads (SARS-CoV-2, n=6; Controls, n=6). Six out of the 12 CBMC samples matched to a PBMCs sample (SARS-CoV-2, n=4; Controls, n=2 Controls). A schematic representation of the study design is illustrated in **Figure 1A**. Clinical and demographic characteristics, and serology results of the cohort are given in **Table S1**. No significant differences were observed between the SARS-CoV-2 and Control groups for the clinical and demographic characteristics except for the serology. Mothers in the SARS-CoV-2 group had a positive SARS-CoV-2 PCR or antigen test before or at the time of inclusion according to the mandatory SmiNet register. Additionally, blood samples from all mothers collected at birth were tested for IgG antibodies against SARS-CoV-2 spike protein and/or nucleocapsid. Mothers with positive antibodies without a prior COVID-19 vaccination registered in the mandatory Swedish vaccination register were classified as infected during pregnancy. COVID-19 vaccination was available to the general population from June 2021, and two mothers who belonged to high-risk groups, one from the SARS-CoV-2 and one from the Control group, were vaccinated for COVID-19, 4 and 5 weeks prior to delivery, respectively. All mothers in the SARS-CoV-2 group had mild COVID-19 without oxygen treatment, and none of them had pregnancy complications, except for one mother in the SARS-CoV-2 group who had preeclampsia. All infants in this sub-study were born between December 2020 and July 2021. None of the infants was born preterm, admitted to a neonatal ward, or had low birthweight.

**Figure 1.**
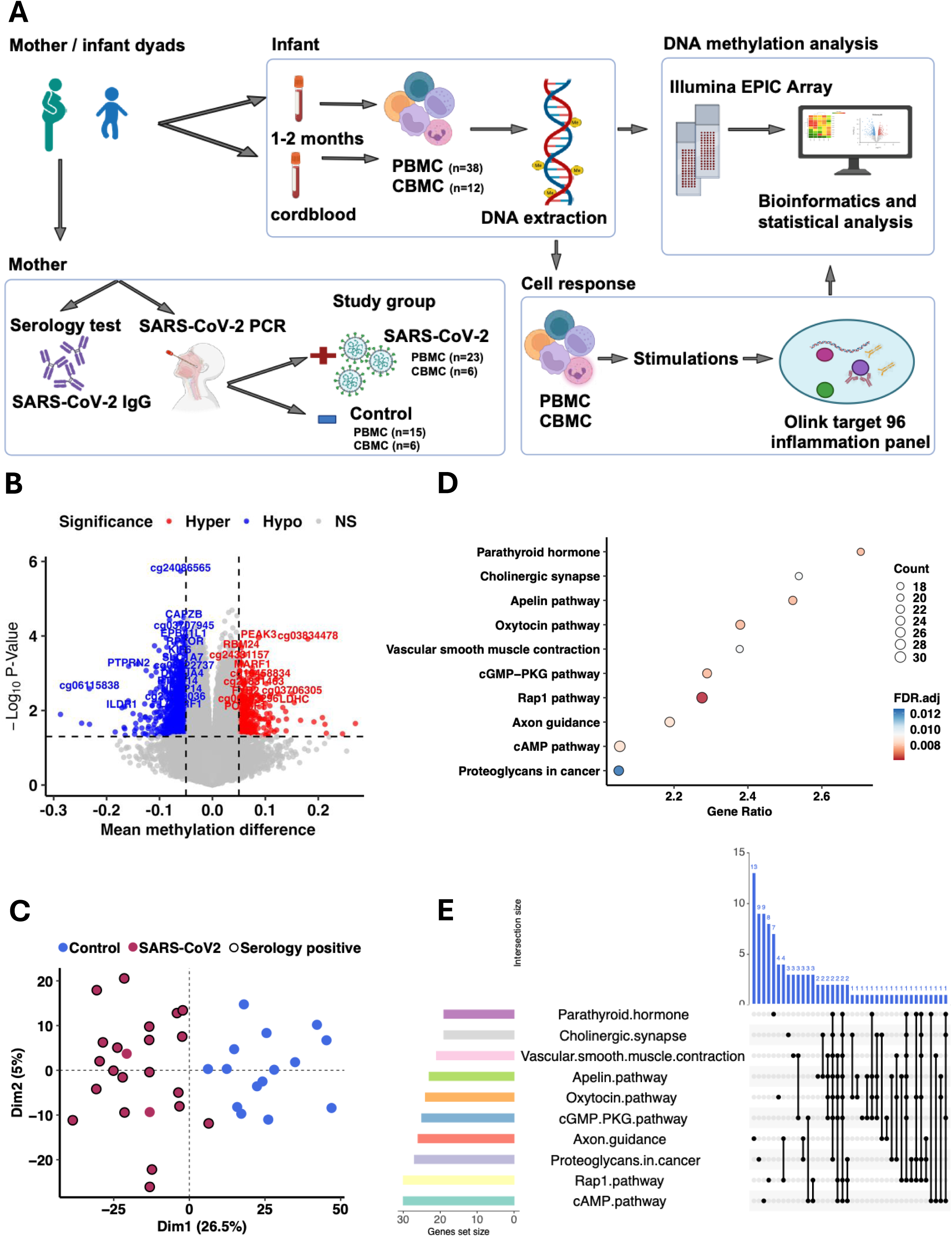
DNA methylome of PBMC samples of infants display intergenerational differences of pregnant women exposed to SARS-CoV-2. **A.** Study overview. Peripheral blood mononuclear cells (PBMCs) were collected from 38 infants at 2 months of age (SARS-CoV-2, n=23; Control, n=15), and cord blood mononuclear cells (CBMCs) were obtained at birth, for a subset of the cohorts (SARS-CoV-2, n=6; Control, n=6). Maternal SARS-CoV-2 exposure during pregnancy was confirmed via PCR and serology. DNA was extracted and DNA methylation profiling was performed using the Illumina EPIC Array, which covers 850K CpG sites. A meta-analysis was performed to identify differences in DNA methylation of genes and their corresponding pathways, associated with maternal SARS-CoV-2 infection. Functional alterations were determined by the assessment of cell responses upon stimulation with SAR-CoV-2 antigens and mitogen using the Olink inflammation 96-panel. **B.** Volcano plot showing the differentially methylated CpGs (DMCs) in PBMCs from the SARS-CoV-2 versus Control group. The dashed horizontal line represents a nominal *p-value* cut-off of 0.05, and the vertical lines represent a cut-off in mean methylation difference > 0.05. Hypermethylated DMCs are shown in red and hypomethylated in blue. **C.** Principal Component analysis of the 2078DMCs showed two distinct clusters separating the SARS-CoV-2 and the Control groups. Serology positive cases are marked with a black outline. **D.** Pathway enrichment analysis of differentially methylated genes (DMGs). Dot plot of the topmost significantly enriched KEGG pathways based on DMGs between groups. The x-axis represents the gene ratio, dot size the gene count, and dot colour the FDR corrected *p-value*. **E.** UpSet plot showing the DMG overlap within the identified KEGG pathways.

### Changes in DNAm patterns in PBMCs following gestational SARS-CoV-2 infection

DNAm data from infants’ PBMCs were analyzed by a cell type deconvolution method to assess cell proportions, revealing no significant differences between the SARS-CoV-2 and the Control groups (**Figure S1A**). Singular value decomposition (SVD) analysis indicated that age and sex contributed significant variation in the DNAm data, and these covariates were therefore included in the linear model to identify differentially methylated CpG sites (DMCs) (**Figure 1B, S1B**). When comparing the PBMC DNAm data from SARS-CoV-2 and Control infants, we identified 2078 DMCs (nominal p*-value* < 0.01) (**Table S2**), of which 833 were hypermethylated and 1245 were hypomethylated, as represented in the volcano plot and a principal component analysis (PCA) plot (**Figure 1B, 1C**). The identified DMCs mapped to 1264 unique differentially methylated genes (DMGs), including 454 hypermethylated, 775 hypomethylated, and 35 with mixed methylation patterns (**Table S3**). In order to investigate the biological significance of the DMGs in infants born to SARS-CoV-2-infected mothers, we performed Kyoto Encyclopedia of Genes and Genomes (KEGG) enrichment analysis. Among the top KEGG-enriched pathways that passed the Benjamini-Hochberg false discovery rate (FDR-corrected *p-value* < 0.05 at 5% significance) were the cholinergic synapse, the apelin signaling pathway, axon guidance, vascular smooth muscle contraction and the production/action of the parathyroid hormone (**Figure 1D and Table S4**). The number of DMGs that overlap between the identified pathways is depicted in the UpSet plot (**Figure 1E**). Pathways with more specific effects included axon guidance, cAMP, Rap1, parathyroid hormone, oxytocin, cholinergic synapse, and apelin, which had unique gene counts of 13, 9, 8, 7, 4, 3, and 2, respectively.

In order to explore whether the trimester of maternal SARS-CoV-2 infection was associated with distinct DNAm patterns in different trimesters, we compared DNAm data from PBMCs of the Control group (n=15) with the group testing positive for SARS-CoV-2 infection in the first trimester (n=5), the second trimester (n=11), and the third trimester (n=7) and identified 86, 84, and 72 DMCs, respectively (nominal *p-value* < 0.05 and mean methylation difference (MMD) > 0.1 except for the second trimester where MMD > 0.05) (**Table S5**). KEGG pathway analysis highlighted trimester-specific changes, including olfactory transduction (first trimester), Notch signalling pathway (second trimester), and the lysosome pathway (third trimester) (**Figure 2A, 2B and 2C**). The beta values of the top four DMCs in each trimester are shown in the boxplots. (**Figure 2D, 2E and 2F**).

**Figure 2.**
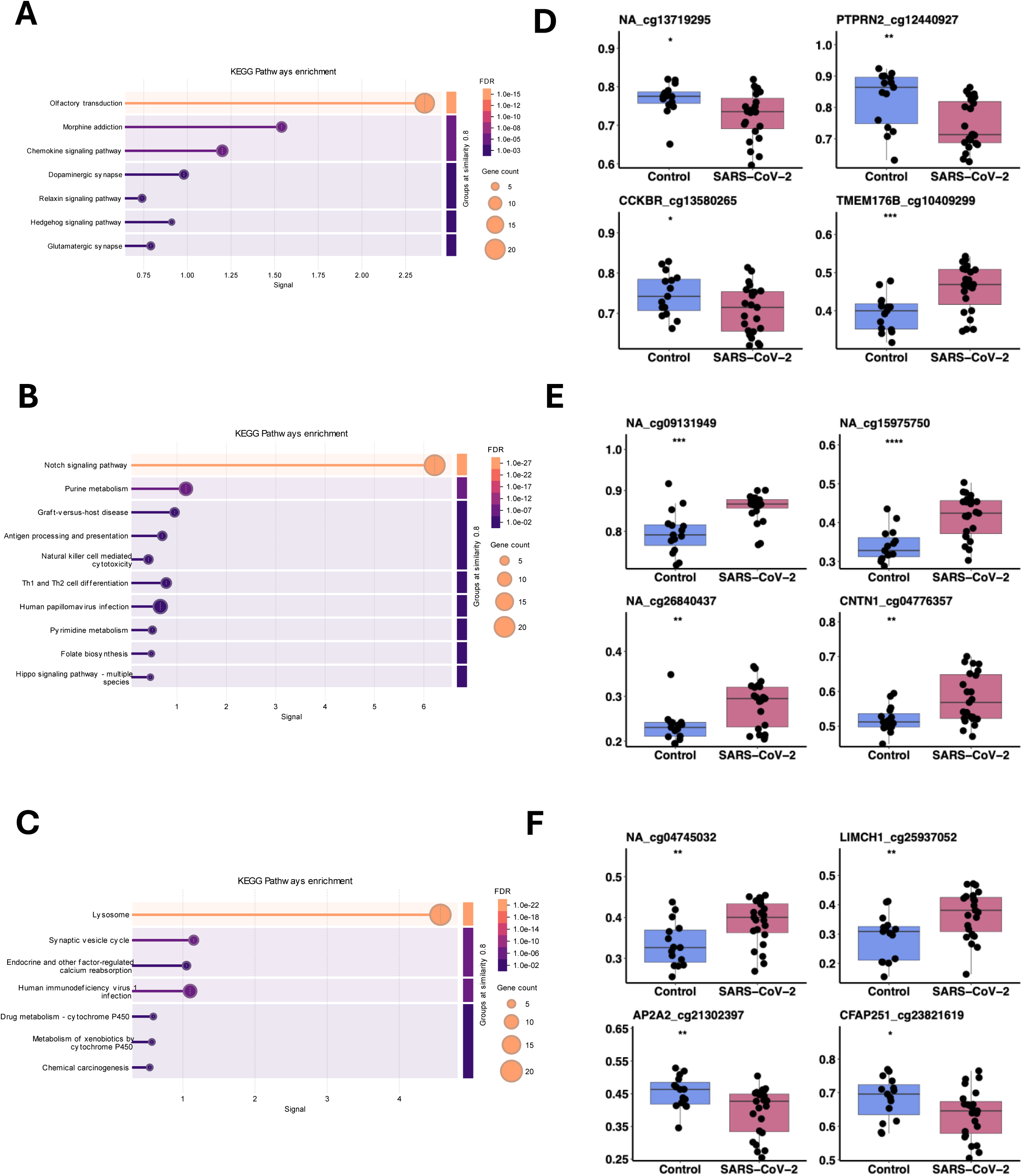
Impact of the trimester of maternal SARS-CoV-2 infection on DNA methylation patterns in infants’ PBMCs. (A-C) KEGG pathways of differentially methylated genes identified in PBMCs based on the trimester during which the mother contracted SARS-CoV-2, first trimester (A) second trimester (B) and third trimester (C). (D–F) Boxplots showing beta values of the top four differentially methylated CpG sites for first, second and third trimester in D, E and F, respectively. Statistical significance was assessed using two-sided unpaired t-tests; significance levels are indicated by asterisks (*).

### DNAm patterns in CBMCs following gestational SARS-CoV-2 infection

We next explored the DNAm patterns in CBMCs at birth between SARS-CoV-2 and Control groups in order to address whether the DNAm patterns in CBMCs overlapped with those identified in PBMCs at 1-2 months. Similarly to PBMCs, cell type deconvolution of cell proportions based on DNAm data showed no significant differences between the SARS-CoV-2 and Control groups in CBMCs (**Figure S2A**). SVD analysis highlighted a significant variation in the DNAm data related to study groups (**Figure S2B**) with 5.3% of the variance explained in the 5^th^ dimension as visualized in the PCA analysis (**Figure S2C**). We identified 894 DMCs mapped to 501 DMGs in CBMCs (nominal *p-value* < 0.05 and MMD > 0.1) (**Table S6 and Figure 3A**), whose beta values revealed a distinct DNAm pattern that separated the groups in a hierarchical cluster analysis (**Figure 3B**). The top 10 KEGG-enriched pathways for the DMGs revealed pathways related to protein and lysine degradation, calcium reabsorption, type-I diabetes, Hippo and Ras1 signaling pathways and adrenergic signaling to cardiomyocytes (**Figure 3C and Table S4**). The number of DMGs that overlap between the identified pathways is depicted in the UpSet plot (**Figure 3D**). The Rap 1 pathway was identified in both CMBCs and PBMCs (**Figure 3D**), while several individual DMGs were shared between the different pathways (e.g. Adrenergic cardiomyocytes and oxytocin pathways; Rap1 and platelet activation; Platelet activation and cGMP/PKG; Platelet activation and vascular smooth muscle contraction). Among the distinct DNAm patterns identified in CBMCs and PBMCs, we identified overlapping changes in 69 genes (**Figure S3A**). The top 10 KEGG-enriched pathways for these 69 genes included those related to O-glycan biosynthesis, various endocrine-related pathways, glutamatergic synapse and platelet activation. (**Figure S3B, and Table S4**).

**Figure 3.**
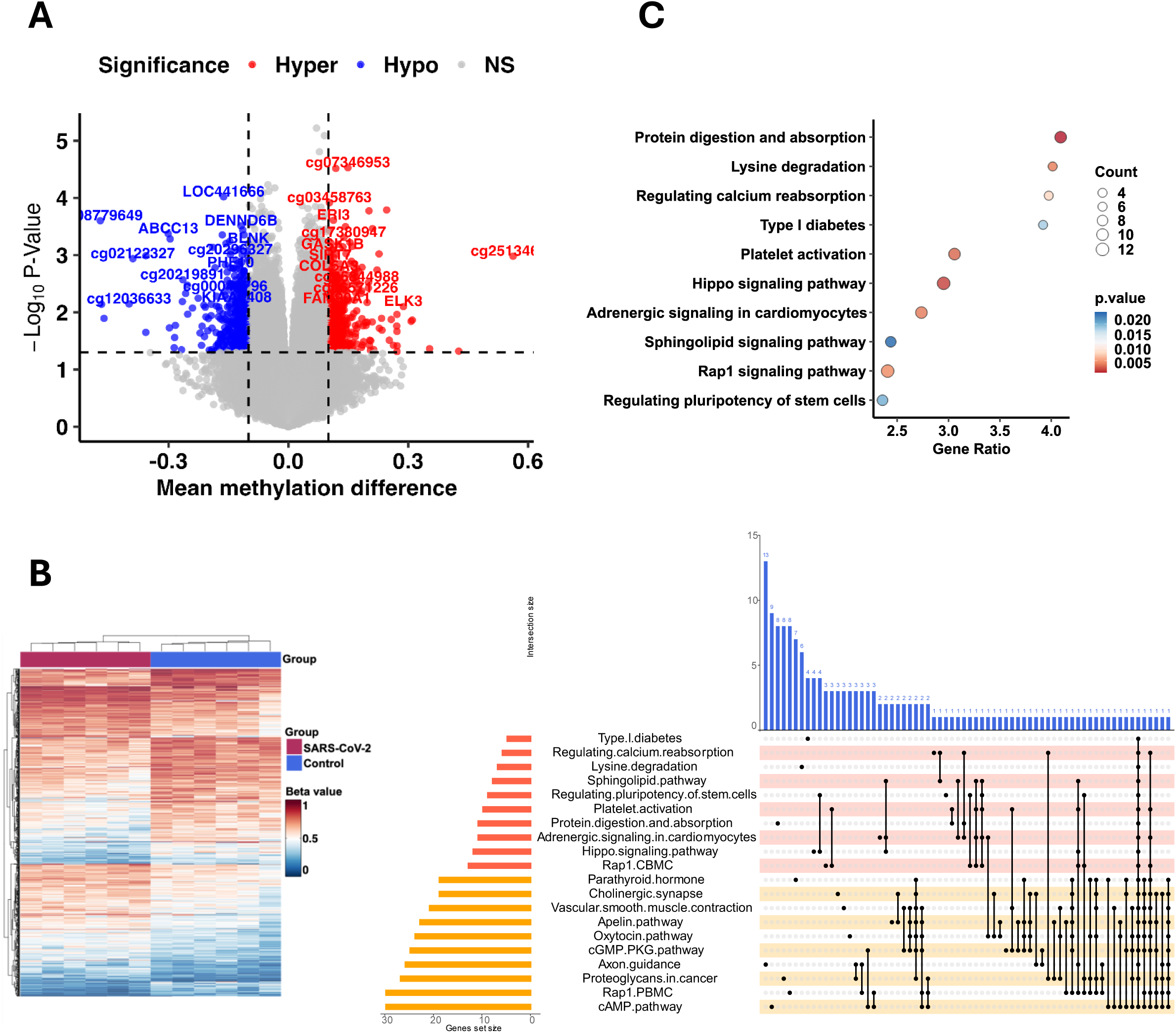
DNA methylation patterns at birth in CBMCs of dyads following gestational SARS-CoV-2 infection. **A.** Volcano plot showing the differentially methylated CpGs (DMCs) in CBMCs from the SARS-CoV-2 versus the Control group. Hypermethylated DMCs are shown in red and hypomethylated in blue. The dashed horizontal line represents a *p-value* cut-off of 0.05, and the vertical lines represent a cut-off in mean methylation difference > 0.1. Hypermethylated DMCs are shown in red and hypomethylated in blue. **B.** Heatmap representing an unsupervised hierarchical clustering analysis of individual beta values for the 894 differentially methylated CpGs (DMCs) between SARS-CoV-2 and Control groups. **C.** Pathway enrichment analysis of differentially methylated genes (DMGs). Dot plot of the 10 most significantly enriched KEGG pathways based on DMGs between SARS-CoV-2 and Control groups. The x-axis represents the gene ratio, dot size the gene count and dot colour the FDR corrected *p-value*. **D.** UpSet plot showing the DMGs overlap between the identified KEGG pathways in both CBMCs (orange) and PBMCs (yellow).

### DNAm changes are correlated to functional alterations in PBMCs which map to SARS-CoV-2-exploited gene products

To validate whether differences in DNAm patterns reflect functional differences, we stimulated PBMCs with two sets of SARS-CoV-2 T cell epitopes, and with a mitogen. Significant differences were noted in mitogen-stimulated samples for AXIN1, CASP8, STAMBP, and OPG/TNFRSF11B with an FDR-corrected *p-value* < 0.05 (**Figure 4A and S4**) here onwards referred as differentially expressed proteins (DEP), whereas six additional proteins passed a nominal *p-value* < 0.05 (**Table S7**). Although a tendency towards dampened expression of these markers was observed in controls of SARS-CoV-2 antigen-stimulated samples, this observation did not reach significance (**Figure 4A)**. No differences were observed in any of the *in vitro* stimulations of CBMCs and therefore not included in the current analysis.

**Figure 4.**
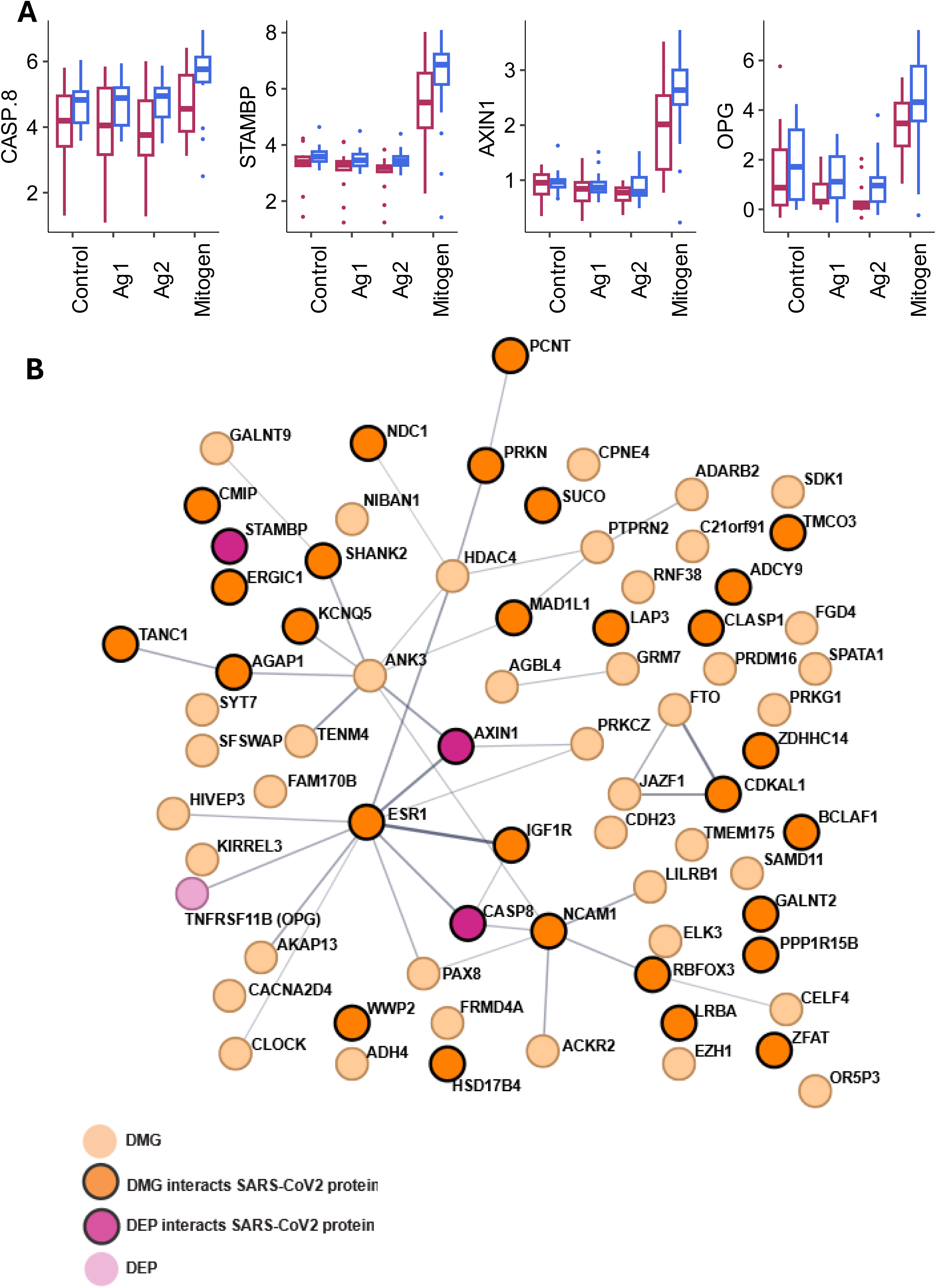
Overlap and functional Insights of differentially methylated genes and differentially expressed proteins in PBMCs and CBMCs. **A.** Boxplot illustrating the statistically significant (FDR corrected *p-value* < 0.05) differentially expressed proteins (DEP) in PBMCs following in vitro stimulation with medium (Control), SARS-CoV-2 peptides (Ag1 and Ag2) and a mitogen between SARS-CoV-2 and Control groups **B.** Protein-protein interaction network showing shared DMGs in PBMCs and CBMCs (orange) and differentially expressed proteins (DEP) from Olink data (purple). Notably, 32 out of the 73 proteins interact with SARS-CoV-2 proteins (see Table S8)

We constructed a protein-protein interaction network for the four DEP that passed the FDR-corrected *p*-value using STRING-db, incorporating the common 69 DMGs from CBMCs and PBMCs. Genes with collective centrality scores in that network were ESR1 (Estrogen receptor-1), ANK3 (Ankyrin 3), NCAM1 (Neural Cell Adhesion Molecule 1), HDAC4 (Histone Deacetylase 4), AXIN1, CASP8 and IGFIR (Insulin Like Growth Factor 1 Receptor) (**Figure 4B**). Out of the total 73 (69 DMG-derived and 4 differentially expressed proteins, DEP) proteins, 32 (highlighted in black outline) have been shown to interact with SARS-CoV-2 proteins (**Figure 4B).** Their corresponding, interacting partners are listed in **Table S8**. The observed interaction (32 out of 73 genes) with the SARS-CoV-2 interactome showed statistically significant overlap (hypergeometric test, p-*value* < 0.001), using a background of human genes included in the Illumina array (n = 22,049) and unique SARS-CoV-2 interactors (n = 5,925).

## Discussion

Previous research on antenatal exposure to infections suggests a mechanistic link between maternal immune activation, reprogramming of the offspring’s epigenome, and subsequent effects on infants.^16^ The COVID-19 pandemic provides the possibility to study this phenomenon due to the large scale of infected pregnant women for the very first time with a specific virus. Inflammatory cytokines induced by maternal immune activation have been suggested to mediate epigenetic changes in the offspring, such as Interleukin-17α, by regulating histone deacetylase activity, and IL-6 by boosting the activity of DNMT1.^17^ Maternal immune activation has been proposed to act as a disease primer, rendering the infants more susceptible to the combined effects of genetic mutations/polymorphisms and subsequent exposures to risk factors in developing neuropsychiatric and neurodegenerative disorders later in life.^13,18^ In the context of gestational COVID-19, the cytokine/chemokine profile of umbilical cord blood samples has demonstrated an upregulation for IL-6, IP-10, and IL-8,^19^ however, whether this contributes to changes in DNAm remains unknown.

In this study, we addressed changes in DNAm in infants born to mothers with asymptomatic or mild gestational SARS-CoV-2 infection compared to infants born to non-infected mothers. The identified DMGs partially overlapped in PBMCs and CBMCs, and DMGs that intersected related to pathways involved in various aspects of different cell adhesion systems (Rap1, proteoglycans in cancer), blood pressure regulation (adrenergic signalling, vascular smooth muscle contraction), and endocrine regulation (oestrogen signalling and parathyroid hormone regulation). In PBMCs, DMGs mapped to pathways related to neurodevelopment (axon guidance, cholinergic synapse, and the apelin pathway), and concurrently, the functional repertoire of some of these pathways extends to multiple biological processes, making interpretation difficult. On the other hand, apelin, an integral hormone in hypothalamo-pituitary axis (HPA) stress-response and in synaptic plasticity,^20^ has a core function in cardiovascular physiology by influencing blood pressure^21^ and cardiac contractility,^22^ and we and others have identified this as differentially methylated in other-COVID-19-repated studies.^23–26^ Of note, apelin is degraded by ACE2, the SARS-CoV-2 entry receptor.^22,27^ Furthermore, recent studies have proposed a link between antenatal COVID-19 and an increased risk of neurodevelopmental and neuropsychiatric manifestations in the offspring.^28^ Infants exposed to antenatal severe/critical COVID-19 display a ten-fold higher frequency of developmental delay compared to pre-pandemic controls.^28^ Higher frequency of neurodevelopmental impairment at 12 months of age was demonstrated, which was accompanied with DNAm changes in buccal swabs and linked to behavioral problems such as autism.^29^ A pilot study with a limited number of buccal swab samples from 3-month-old infants showed methylation changes enriched in genes related to neurodevelopmental pathways,^30^ similar to our findings in PBMCs. Conversely, DNAm patterns in the maternal-fetal interface (placental cells) in the third trimester^31^ and cord blood cells at birth^23^ after COVID-19, have showed broader effects on the epigenome that related to aberrant immune activation and angiogenesis, cardiac, renal, hepatic, neurological diseases, developmental and immunological disorders.

The association between the gestational window of viral infections and child fetal neurodevelopment, learning, behavioral and psychiatric disorders has been a subject of investigation.^32,33^ In our study, DMGs in infants born to mother with SARS-CoV-2-infection during the first trimester mapped to pathways related to olfactory transduction, similarly to adults following mild COVID-19,^4^ whereas second-trimester infection associated to involvement of Notch signaling, a vital pathway in the development and function of the neuronal, cardiovascular and endocrine systems^34^ and immune-related pathways. In animal studies, injection of the viral analogue poly I:C during early or late pregnancy in mice resulted in distinct DNA methylation signatures in progeny brain regions^35^ and, in rats, resulted in different expression patterns of brain-associated miRNAs.^36^ Antenatal SARS-CoV-2 infection in the first and second trimester has been proposed to have an impact on birth outcomes, such as a higher risk for stillbirth and lower gestational age at delivery, regardless of maternal infection severity.^37^ The biological significance of the gestational window of SARS-CoV-2 infection and the observed change in the epigenetic patterns requires further studies with larger cohorts to be determined.

The functional assessment of PBMC responses to a non-pathogen specific stimulus (mitogen) detected reduced expression for AXIN1, CASP8, STAMBP, and OPG. Overall, functional differences in PBMCs point towards modulation of cell death processes like apoptosis and necroptosis. An intriguing aspect of our findings is that among the identified DMGs, we found 32 genes whose gene products interact with SARS-CoV-2 proteins, including AXIN1, CASP8, and STAMBP. The observed lack of significant PBMC response to SARS-specific epitopes further indicates that the immune system of the fetus never encountered the SARS-CoV-2 virus during pregnancy.

The strength of the current study is that it included mother-infant dyads in the early phase of the pandemic, with most mothers not yet being vaccinated or previously exposed to SARS-CoV-2, and that the infections were mild. Samples from both the SARS-CoV-2 and the Control group were collected at the same period, which reduces the risk of confounders associated with studies using historical controls. The diagnosis of SARS-CoV-2 infection was confirmed both with nasopharyngeal swab PCR and serology in blood, and the DNAm analyses were complemented with functional analyses of PBMCs and CBMCs. A limitation of our study is the small number of samples for CBMCs and following sample stratification according to trimester, that may impose a power issue, hence nominal p-values were used instead in this case. The analyses were conducted in PBMCs, consequently epigenetic changes in other body compartments were not covered. However, several studies have identified DNAm changes in blood samples mapping to pathways of strong relevance for diseases in distal organs, highlighting the usefulness of blood-based DNAm analyses as proxy biomarkers. For example, epigenetic modifications in pathways relevant for kidney function were found in the blood of patients with chronic kidney disease.^38^ Furthermore, a restricted panel of inflammation-related proteins was employed for the analyses of *in vitro* assays, which may limit the overlap of DMGs and DEPs and constrain the functional interpretation of the findings. Lastly, we lack analysis of inflammatory cytokines in blood that could potentially correlate with the observed DNAm changes.

### Conclusions

Maternal COVID-19 during pregnancy, even in its mild form, was associated with alterations in the DNA methylome in the progeny. The pathways identified are primarily involved in neurodevelopment and blood pressure regulation, while several of their corresponding genes are known to act also as viral-interacting factors. The global impact of those priming effects is not yet known, nor is their persistence over time. However, the identified overlap of these targets with processes regulating fundamental developmental processes such as the formation of the nervous system points toward a possible cost-benefit relationship of detrimental consequences as a cost for improved infection resilience. Further mechanistic studies are needed to clarify this relationship.

## Key resource table

Information about the key resource materials is provided on the key resource table (xlsx).

## Data Availability

The datasets generated during the current study are not publicly available due to ethical dilemmas in traceability of DNA methylation data, but processed data that is pseudonymized and depleted of genetic variant information (beta matrixes with Illumina probe IDs and beta values) will be available upon request through the Federated European Genome-phenome Archive (FEGA) Sweden controlled-access repository, upon publication. Bioinformatic pipelines used to analyze the data and to generate graphs and figures will be available on the following GitHub account upon publication:
(https://github.com/Lerm-Lab/COPE_epigenetics/).

## Resource availability

### Lead contact

Requests for further information and resources should be directed to and will be fulfilled by the lead contact, Maria Lerm.

### Data and code availability

The datasets generated during the current study are not publicly available due to ethical dilemmas in traceability of DNA methylation data, but processed data that is pseudonymized and depleted of genetic variant information (beta matrixes with Illumina probe IDs and beta values) will be available upon request through the Federated European Genome-phenome Archive (FEGA) Sweden controlled-access repository, upon publication. Bioinformatic pipelines used to analyze the data and to generate graphs and figures will be available on the following GitHub account upon publication: (https://github.com/Lerm-Lab/COPE_epigenetics/).

## Acknowledgments

We would like to thank all families participating in this study as well as the study nurses helping us with the clinical follow-up. Moreover, we want to acknowledge SciLifeLab Autoimmunity and Serology Profiling unit, Stockholm, for the serology analysis, SciLifeLab affinity proteomics, Stockholm, for the Olink analysis, Clinical Genomics Linköping, Science for Life Laboratory, Linköping University and Core facility at Medical Faculty, Linköping University for generating the DNA methylation data. We also thank OMDA MedSciNet for helping to maintain the COPE database. We are grateful for the support of the Swedish Network for National Clinical Studies in Obstetrics and Gynaecology, SNAKS (www.snaks.se) and Clinical Studies Sweden in setting up the COPE study. This study was conducted using professional biobank services from Biobank Sweden (https://biobanksverige.se/). The authors would like to acknowledge the support of the SciLifeLab Autoimmunity and Serology Profiling Unit in Solna, Sweden, for performing the serological analyses.

## Funding

The initiation of the COPE study was financed by the Simons Foundation Autism Research Initiative, SFARI (V.S., 863675), Jane and Dan Olsson Foundation (V.S., 2021-02), Stiftelsen Erik & Lily Philipsons minnesfond (V.S., dnr 98) and Swedish Research Council grants (H.B., 2018-00470; S.B.W., 2016-00526). This substudy of the COPE study was supported by Swedish Heart Lung Foundation (M.L., 20220034), Swedish Research Council (A.R., 4.3-2019-00201 & GD-2020-138; T.A., 2020-01111; M.L., 2021-03183 & 2022-01213); Johanna Cocozza foundation (T.A.), Swedish Cancer Society (A.R., 211832Pj01H2/Infection-Autoimmunity-B-lymphoma grant) and local Linköping University funds.

## Author contributions

Conceptualization, M.L. and T.A.; Methodology, E.A., S.S., C.J.; Investigation, E.A., S.S., M.B., C.J., N.Y.**-**F., H.Ö., A.S., H.B., M.V., M.Z., S.B.W., Y.C., M.D., F.A., S.A., O.A., L.B., V.S., M.L., T.A.; Formal Analysis, S.S., E.A.; Data curation, S.S., E.A., S.A., V.S.; Software, S.S.; Visualization, S.S., E.A., M.L.; Writing – Original Draft, E.A., S.S., M.L., T.A.; Writing –Review & Editing, E.A., S.S., M.B., C.J., N.Y.**-**F., H.Ö., A.S., H.B., M.V., M.Z., S.B.W., Y.C., M.D., F.A., S.A., O.A., L.B., V.S., A.R., M.L., T.A. ; Funding Acquisition, A.R., M.L., V.S., H.B., S.B.W., T.A.; Resources, M.L., T.A., V.S.; Supervision, M.L., T.A., A.R.; Project administration, M.L., T.A., V.S., S.A.

## Declaration of interests

M.L. and S.S. are founders of a company, which has the ambition to develop DNAm-based biomarkers for a variety of conditions, including Post-COVID Conditions. The other authors declare no competing interests.

## Declaration of generative AI and AI-assisted technologies

The ChatGPT was used for language improvement.

## STAR Methods

### Study population

This research was conducted as a sub-project within the larger COVID-19 in Pregnancy and Early Childhood (COPE) study — a nationwide, multi-center initiative in Sweden that recruited pregnant participants during the COVID-19 pandemic. The COPE study is officially registered on ClinicalTrials.gov (NCT 04433364), and comprehensive details of its methodology are available in a previously published protocol. ^15^ The COPE dataset is linked to Swedish health and quality registers and includes a biobank (registration number: Biobank Väst, ID 890). Briefly, all women, aged 18 years or older, with a viable singleton gestation pregnancy were eligible for this substudy. Recruitment of pregnant women occurred at different time points during the pregnancy, for example, at the first- or second-trimester ultrasound screening visit, on admission for pregnancy complications or admission to the delivery units. To be eligible for the SARS-CoV-2 group, mothers had to have a SARS-CoV-2 infection at any point during pregnancy as documented by 1) nasopharyngeal swab PCR testing in the Swedish Register for Mandatory Registration for notifiable Diseases (SmiNet) or 2) one of the following ICD-10 codes in any of the included registers: U07.1; U07.2. or 3) positive SARS-CoV-2 IgG antibodies during pregnancy or at delivery without prior vaccination. The control group consisted of women without any positive test for SARS-CoV-2 infection during the current pregnancy and without U07.1 or U07.2 diagnosis and with no positive SARS-CoV-2 IgG antibodies during pregnancy or at delivery or positive SARS-CoV-2 IgG antibodies during pregnancy or at delivery due to prior vaccination. The present sub-study included only mothers to offsprings from which peripheral blood mononuclear cells (PBMC) were isolated from blood samples and only from one of the participating study sites (Linköping). Participating women and their partners received oral and written information about the study and provided written consent. The study was approved by Swedish Ethical Review Authority (dnr 2020-02189 and amendments 2020-02848, 2020-05016, 2020-06696, and 2021-00870) and received national biobank approval by the Biobank Väst (dnr B2000526:970).

### Serology analysis for SARS-CoV-2 antibodies in maternal blood

EDTA plasma was collected at inclusion during pregnancy, at birth and 1-2 months postpartum from the mothers. A Luminex-based assay was employed for the serology test of IgG antibodies against two SARS-CoV-2 spike protein antigens (a soluble trimeric domain, and the S1 domain of the spike glycoprotein) and one antigen derived from the C-terminal domain of the nucleocapsid. Positivity in any two of the three antigens defined a positive sample. The analysis was performed by the SciLifeLab Autoimmunity and Serology Profiling unit in Stockholm and is described in detail previously.^39^

### PBMC isolation from 1-2 months old infants and cord blood

Infant venous blood was drawn into heparin treated tubes (Vacuette, Greiner Bio-One GmbH, Austria) and cord blood was collected by cutting the umbilical cord at the placental site, carefully cleaning it to avoid contamination of mother’s blood and then collecting into heparin treated tubes (Vacuette). Peripheral blood mononuclear cells (PBMCs) were isolated using Ficoll-Paque density gradient (Cytiva, Uppsala, Sweden), washed three times in RPMI 1640 medium (Gibco, Fisher Scientific, Sweden) supplemented with 2% heat inactivated FCS (Cytiva). Cells were frozen in 10% dimethyl sulfoxide (DMSO, Merck), 40% RPMI 1640 and 50% FCS and kept at -150 °C until further processing.

After thawing, the cells were washed twice in cell culture medium (RPMI medium 1640, 31870-025, 10% fetal bovine serum, 1% penicillin/streptomycin, 15140, 1% L-glutamine, 25030081, all from Gibco, Fisher Scientific, Sweden) further on termed as complete culture medium, prior to further processing.

### Epigenetic data from whole blood and cord blood PBMCs

DNA was extracted from PBMC using QIAamp DNA Mini Kit on the QIAcube Connect platform (Qiagen, 51304, 9002864, Germany). DNA quantity was measured with Qubit HS dsDNA (Q32851, Invitrogen, USA) and the quality was evaluated with with NanoDrop Spectrophotometer (ND-1000, Thermo Fisher Scientific, USA).

The methylation array data was generated by Clinical Genomics Linköping, Linköping University, Sweden. For each sample, 250 ng of DNA extracted from PBMCs was subjected to bisulfite conversion using EZ DNA Methylation Kit (Zymo Research) and further processed according to the Illumina Infinium MethylationEPIC v.1.0 (850K) protocol. The BeadChip arrays were scanned on the NextSeq 550 (Illumina) instrument.

### DNA methylation analyses

The raw IDAT files were loaded and analyzed in R v4.1.3 using ChAMP package v2.24.0.^40^ The DNAm data was pre-processed using a robust filtering criterion by removing the probes (i) with a detection *p-value* > 0.05, (ii) on sex chromosome to get rid of any gender biases and as female X-inactivation skews the distribution of beta values, (iii) that are cross-reactive, (iv) containing known single nucleotide polymorphisms (SNPs). After filtering, we performed the quality control.

We next performed the beta-mixture quantile normalization (BMIQ) to adjust for the inherently different designs of the Illumina DNA methylation arrays.^41^ The beta-values and M-values of the samples were calculated against each probe per sample. Thereafter, we performed singular value decomposition (SVD) analysis to identify the most significant components of variation within the filtered normalized DNAm data. Any technical bias or confounders could be identified using SVD. Batch corrections were performed in the PBMCs data for the technical factors/artifacts such as slide and array using ComBat from the SVA package v3.42.0^42^ followed by SVD analysis on the corrected data. The SVD analysis of CBMCs data did not show any variation due to slide and array therefore no batch correction was applied.

To identify the possible differences in cellular content of each sample, the cell type proportions in the samples were calculated with the Epigenetic Dissection of Intra-Sample-Heterogeneity (EpiDISH) package v2.^1^5.1 in R.^43^ It is based on Houseman algorithm using the whole blood reference of 12 blood cell subtypes “cent12CT.m” (i.e. NK cells, monocytes, Treg, Basophils as well as memory and naive cells for B cells, CD4T and CD8T cells) for EPIC array.^44,45^ The quality of the filtered, normalized and batch corrected data is checked using the different metrices such as MDS plot, hierarchical clustering and density plot of beta distribution. Principle component analysis (PCA) was performed using the packages FactoMineR and factoextra.^46^

### Differential DNAm analysis

To identify the DMCs between the SARS-CoV-2 and Control groups, we used limma package v3.50.3 in R.^47^ A linear model was fitted to the filtered, normalized and corrected DNAm data. We identified age and sex as sources of variation that were still present after SVD correction and were therefore, added as co-variates in the linear model (lmFit). Moderated t-statistics, log2 fold change (referred to as mean methylation difference, MMD), nominal *p-value* and FDR corrected *p-value* were computed for each probe. None of the DMCs passed through the multiple testing. We defined the significant DMCs between the groups (SARS-CoV-2 vs Control) with the cutoffs nominal *p-value* < 0.01 criteria in PBMCs infants and a cutoff of MMD > 0.1 and a nominal *p-value* < 0.05 criteria in cord blood DNAm data. Heatmaps were generated using ComplexHeatmap package v2.10.0 and dendsort v0.3.4.^48,49^ The identified DMCs were mapped to their corresponding genes (DMGs) using Illumina methylation annotations for EPIC array *ilm10b4.hg19*.

### Pathway enrichment analysis

Next, the identified DMGs were assessed using Kyoto Encyclopedia of Genes and Genomes (KEGG) pathway.^50^ Pathway over-representation analysis was performed using WEB-based GEneSeT AnaLysis Toolkit (WebGestalt) with KEGG as functional database and default settings for analysis.^51^ To enhance the visualization and better understanding of the enrichment results, ggplot2^52^ v3.4.4 was used to generate dotplot. Genes intersection in different pathways were plotted with UpsetR package v1.4.0.^53^

### Overlap of DMGs and DEPs with the SARS-CoV-2 Interactome

To identify proteins interacting with the SARS-CoV-2 interactome, we utilized publicly available SARS-CoV-2 interacting proteins and human genes curated in BioGRID (v4.4.210, www.biogrid.org).^54^ We analyzed 60 differentially methylated genes (DMGs) shared between PBMCs and CBMCs, along with three differentially expressed proteins (DEPs), against the SARS-CoV-2 interactome from BioGRID, identifying 26 overlapping genes.

To evaluate the enrichment of DMGs for SARS-CoV-2 interactors, we performed a hypergeometric test using the human genes from the Illumina array (n = 22,049) as background and unique SARS-CoV-2 interactors (n = 5,925). These 63 overlapping genes served as seed genes for constructing a protein-protein interaction (PPI) network in STRING-db (confidence score > 0.4).^55^ The resulting PPI network was visualized using Cytoscape v3.8.0.^56^

### Cell responses of stimulated PBMCs

The QuantiFERON™ SARS-CoV-2 assay (QFN) (Qiagen, Hilden, Germany) was performed in isolated PBMCs from all newborns (n=31) and 12 CBMCs, 6 matched and 6 non/matched. The assay included four collection tubes: Nil tube, as negative control for background subtraction; Ag1 tube, for stimulation of Spike1-specific CD4+ T cells; Ag2 tube, for stimulation of Spike1- and Spike2-specific CD4+ and CD8+ T cells; and Mitogen tube, as positive control. 1×10^6^ PBMCs or CBMCs in 1 ml of complete culture medium were incubated in each tube at 37°C, 5% CO_2_ for 6 days. Following incubation, the tubes were centrifuged, and supernatants were collected and analyzed for IFN-γ release according to the manufacturer’s protocol. Results were expressed in international units per milliliter (IU/mL). Nil values were subtracted from Ag1 and Ag2 values to obtain final adjusted IFN-γ concentrations. A QuantiFERON-reactive status was defined when, in Ag1 and/or Ag2 tubes, an IFN-γ concentration of at least 0.15 IU/ml was detectable.

Supernatants from stimulated PBMCs (n=32, 16 from the SARS-CoV-2 and 16 from the Control group) and from stimulated CBMCs (n=12) were used for protein expression analysis by commercially available multiplex proximity extension assay from Olink proteomics and performed in SciLifeLab in Stockholm. Protein expression was measured for each stimulation (Nil, Ag1, Ag2 and Mitogen) using the Olink Target 96 Inflammation panel (v.3025).

Protein levels were expressed in a log2 scale as normalized protein expression values (NPX). The analysis of proteomic data was performed using the OlinkAnalyzer v3.7.0 and a custom script in R.^57^ Preprocessing of the data included the removal of the assays in which the expression levels were below the limit of detection (LOD) in >75% of samples. SVD analysis was used to find the confounders and covariates and was considered in the model as explained above. The baseline-corrected NPX values of remaining 62 assays were analyzed using two-way ANOVA and Post Hoc test to find the differentially expressed proteins (DEPs) with an FDR-corrected *p-value* < 0.05.

### Statistics

All statistical analysis was performed in R v4.1.3 and bioconductor packages. Initial descriptive analysis of demographic variables was performed on the available information about the mother (age, parity, BMI, country of origin, comorbidities, mode of birth, SARS-CoV-2-positive serology IgG test) and of the infants (sex, gestational age at birth, and birthweight) (**Table S1**). Continuous variables were tested for normality using the Shapiro Wilk test followed by non-parametric Mann-Whitney U test. Categorial variables were analyzed using the Chi-squared test. To study the differences in cell type proportions between the groups, we used the Kruskal Wallis test. Multivariate analysis was performed using SVD and PCA techniques to assess the association of DNAm and variables. For differential methylation/protein analysis, the data was fitted to a linear model. A two-way ANOVA and Post Hoc test were further used to find the DEPs. *P-values* were adjusted for multiple testing using the Benjamini-Hochberg procedure for False Discovery Rate (FDR) correction at 5%. In case no significant FDR was reached, we used nominal *p-value* < 0.05. To assess the enrichment of DMGs for SARS-CoV-2 interactors, we performed a hypergeometric test.

## Declaration of generative AI and AI-assisted technologies in the writing process

During the preparation of this work the authors used ChatGPT in order to improve the text language. After using this tool/service, the authors reviewed and edited the content as needed and took full responsibility for the content of the publication.

## Notes

### Author Declarations

The Swedish Ethical Review Authority (dnr 2020-02189 and amendments 2020-02848, 2020-05016, 2020-06696, and 2021-00870) gave ethical approval for this work and received national biobank approval by the Biobank Väst (dnr B2000526:970).

