## Supplementary FigureS1 for "DNA methylation changes in infants of mothers with SARS-CoV-2 infection during pregnancy"

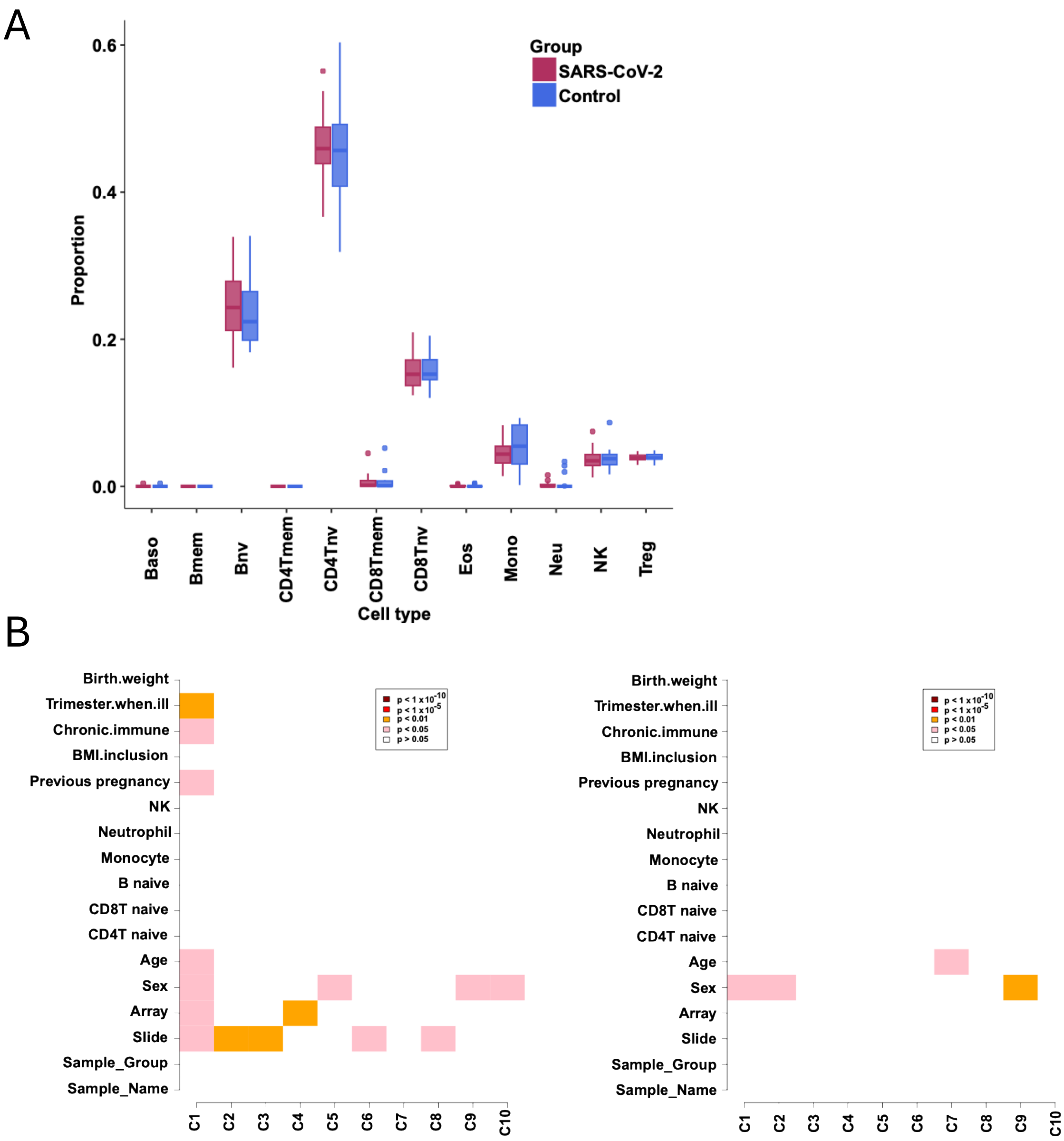


**Supplementary Figure S1. DNA methylation analysis in PBMCs.**
**A.** Relative frequencies (%) of monocytes (mono), neutrophils (neutro), T regulatory cells (Tregs) natural killer cells (NK), CD4 naïve T lymphocytes (CD4Tnv), CD4 memory T lymphocytes (CD4Tmem), CD8 T naïve lymphocytes (CD8Tnv), CD8 T memory lymphocytes (CD8Tmem), B naïve lymphocytes (Bnv), B memory lymphocytes (Bmem), eosinophils (eos), and basophils (baso) based on cell type deconvolution analysis of DNAm data infants' PBMCs from the SARS-CoV-2 (red) and the Control group (blue).
**B.** Singular value decomposition analysis before and after batch correction.
