## Supplementary FigureS2 for "DNA methylation changes in infants of mothers with SARS-CoV-2 infection during pregnancy"

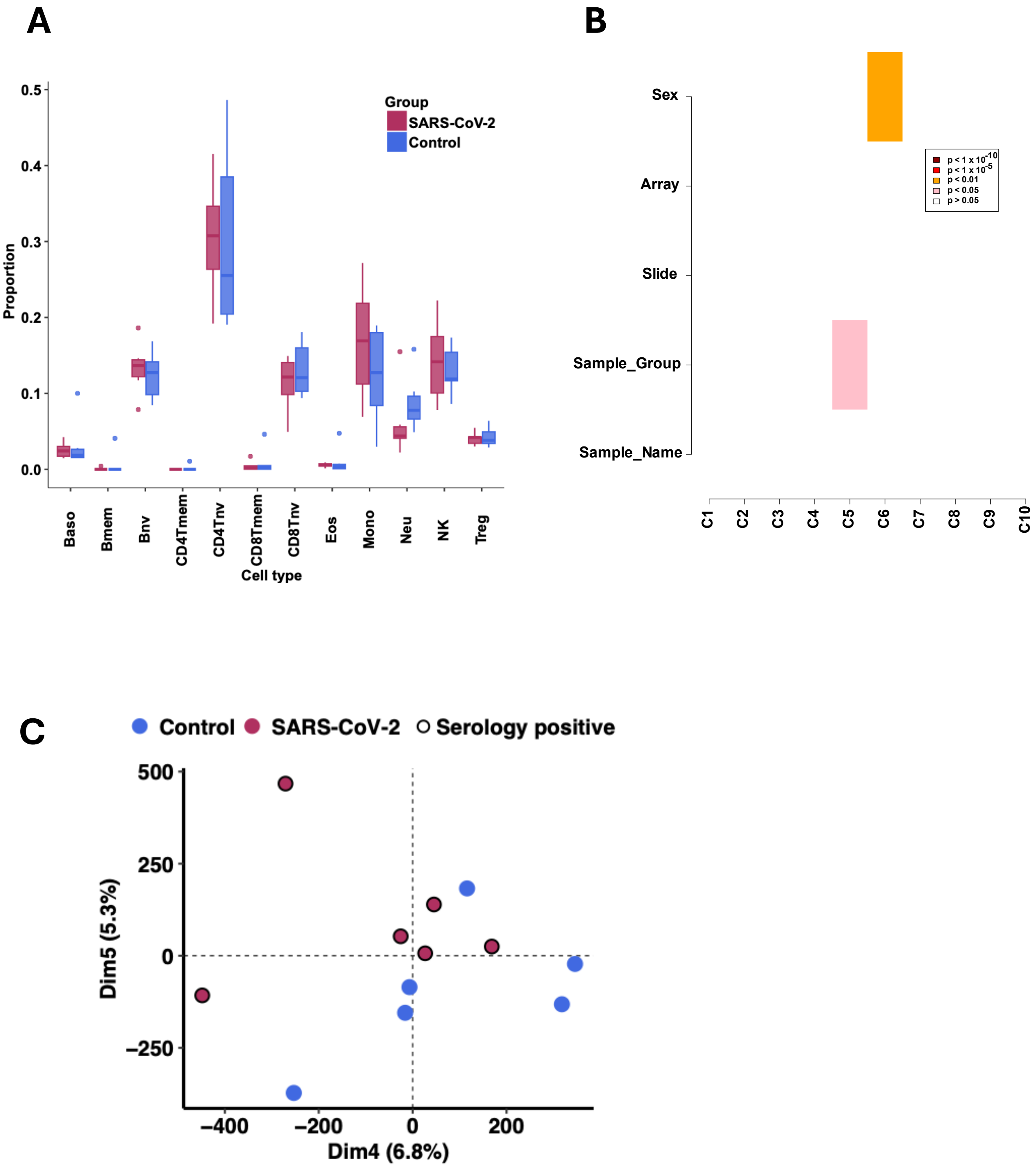


**Supplementary Figure S2. DNA methylation analysis in CBMCs.**

**A.** Relative frequencies (%) of monocytes (mono), neutrophils (neutro), T regulatory cells (Tregs) natural killer cells (NK), CD4 naïve T lymphocytes (CD4Tnv), CD4 memory T lymphocytes (CD4Tmem), CD8 T naïve lymphocytes (CD8Tnv), CD8 T memory lymphocytes (CD8Tmem), B naïve lymphocytes (Bnv), B memory lymphocytes (Bmem), eosinophils (eos), and basophils (baso) based on cell type deconvolution analysis of DNAm data CBMCs from the SARS-CoV-2 (red) and the Control group (blue).

**B.** Singular value decomposition analysis.

**C.** Principal Component analysis of the DNA methylation data at birth in CBMCs of dyads following gestational SARS-CoV-2 infection shows two distinct clusters separating the SARS-CoV-2 vs Control group. Serology positive cases are shown with black outline.
