## Supplementary FigureS3 for "DNA methylation changes in infants of mothers with SARS-CoV-2 infection during pregnancy"

A

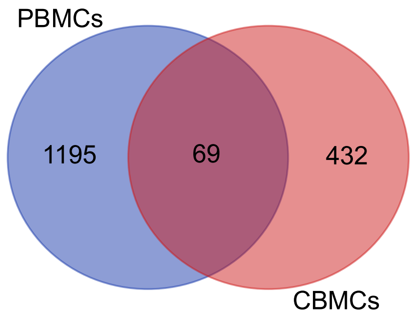


B

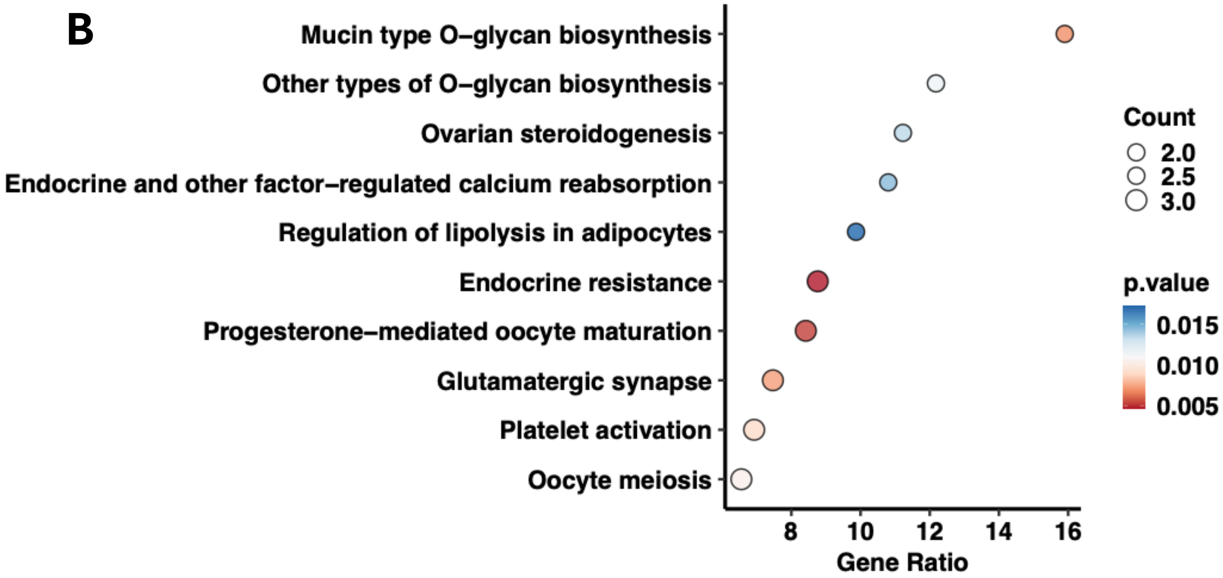


**Supplementary Figure S3. KEGG-enriched pathways of the overlapping differentially methylated genes (DMGs) in CBMCs and PBMCs.**

**A.** Venn diagram illustrating the overlap of differentially methylated genes (DMGs) between PBMCs and CBMCs.

**B.** Pathway enrichment analysis of the sixty-nine differentially methylated genes (DMGs) in CBMCs and PBMCs. Dot plot of the 10 most significantly enriched KEGG pathways based on DMGs between SARS-CoV-2 and Control groups. The x-axis represents the gene ratio, dot size the gene count and dot colour the nominal *p-value* $< 0.05$.
