## Supplementary FigureS4 for "DNA methylation changes in infants of mothers with SARS-CoV-2 infection during pregnancy"

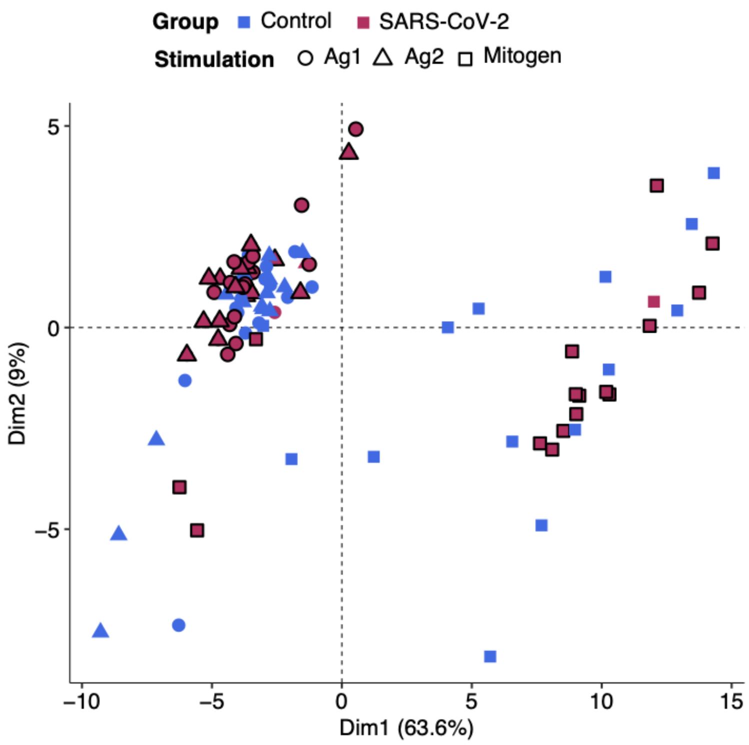


**Supplementary Figure S4. Cell responses of stimulated PBMCs analyzed using Olink data.**

Principal component analysis of baseline-corrected normalized protein expression (NPX) data from PBMCs showing responses to different stimulations.
